# COVID-19 Trends at The University of Tennessee: Predictive Insights from Raw Sewage SARS-CoV-2 Detection and Evaluation and PMMoV as an Indicator for Human Waste

**DOI:** 10.1101/2024.02.03.24302256

**Authors:** Y. Li, K. T. Ash, I. Alamilla, D. C. Joyner, D. E. Williams, P. J. McKay, B. M. Green, S. E. DeBlander, C. M. North, F. Kara-Murdoch, C. M. Swift, T. C. Hazen

## Abstract

Wastewater-based epidemiology (WBE) has become a valuable tool for monitoring the prevalence of SARS-CoV-2 on university campuses. However, concerns about effectiveness of raw sewage as a COVID-19 early warning system still exist, and it’s not clear how useful normalization by simultaneous comparison of Pepper Mild Mottle Virus (PMMoV) is in addressing variations resulting from fecal discharge dilution. This study aims to contribute insights into these aspects by conducting an academic-year field trial at the student residences on the University of Tennessee, Knoxville campus, raw sewage. This was done to investigate the correlations between SARS-CoV-2 concentrations, both with and without PMMoV normalization, and various parameters, including active COVID-19 cases, self-isolations, and their combination among all student residents. Significant positive correlations between SARS-CoV-2 concentrations a week prior, during the monitoring week, and the subsequent week with active cases. Despite these correlations, normalization by PMMoV does not enhance these associations. These findings suggest the potential utility of SARS-CoV-2 concentrations as an early warning indicator and provide valuable insights into the application and limitations of WBE for COVID-19 surveillance, specifically within the context of raw sewage on university campuses.

## Introduction

Wastewater-Based Epidemiology (WBE) emerges as a valuable tool for the early detection and surveillance of COVID-19 outbreaks within university communities. As of December 22, 2023, a study by researchers at the University of California, Merced, reveals that 289 universities across 72 countries, encompassing 4648 sites, have adopted WBE practices (Collaborative 2023). Through routine analysis of wastewater samples from dormitories and other campus facilities, health authorities can promptly identify potential infection clusters and implement preventive measures. This proactive approach curtails virus transmission among students, facilitating swift isolation and contact tracing, ultimately mitigating the impact of outbreaks on campus life.

The significant early warning capability of Wastewater-Based Epidemiology (WBE) is evident in COVID-19 surveillance, encompassing two crucial dimensions. Firstly, it signals the onset of an outbreak at its initial stage, and secondly, it forecasts an impending surge in the number of infected individuals. Noteworthy studies highlight WBE’s effectiveness in providing advanced indications of COVID-19 outbreaks, with reported early warning days varying from two days to three weeks for early COVID-19 trend detection. Karthikeyan et al. (2021) demonstrated accurate predictions of cases by a week and intermediate accuracy at three weeks using city wastewater. Similarly, Randazzo et al. (2020) positive SARS-CoV-2 results were identified in wastewater samples twelve to sixteen days before the official reporting of COVID-19 cases in three out of seven wastewater catchment regions. Medema et al. (2020) identified a similar pattern, with wastewater samples indicating the presence of SARS-CoV-2 six days before the official reporting of the first cases. This detection was explicitly associated with the N3 gene, excluding the N1, N2, and E genes. Nemudryi et al. (2020) found SARS-CoV-2 RNA levels in wastewater exhibiting a lead time of two to four days before clinical results, and they were indicative of symptom onset by five to eight days. Ahmed et al. (2020) emphasized the potential of wastewater monitoring as an early warning system, providing insights into the broader circulation of SARS-CoV-2, particularly in individuals with mild or no symptoms. However, Gerrity et al. (2021) propose that while wastewater surveillance may effectively function as a leading indicator for COVID-19 outbreaks, it may lag as an indicator for declining infection rates due to prolonged viral shedding. This introduces uncertainty concerning the timing and dynamics of infection rate trends as reflected in wastewater data. Moreover, most of the studies primarily examined wastewater rather than raw sewage when making early warning predictions. Additionally, Li et al. (2021) summarized that various methodological challenges can affect the accuracy of prevalence estimation in WBE.

One of the primary challenges in handling SARS-CoV-2 data revolves around the crucial normalization process. Raw sewage, which comprises a diverse range of liquids originating from toilets, sinks, showers, and dishwashing within a building, forms a complex matrix containing human waste components such as feces, urine, sputum, and nasal discharge. The viral levels present in sewage exhibit variability dependent on water usage patterns. To comprehensively understand such sewage, measurements of indicators associated with human waste or fluids are conducted simultaneously with the assessment of SARS-CoV-2 concentrations. Pepper Mild Mottle Virus (PMMoV) is systematically assessed with wastewater due to its acknowledged stability and minimal concentration variation over time ((Li et al. 2023, Li et al. 2023). This virus has been employed to detect pathogenic enteric viruses, as heightened concentrations of PMMoV often indicate increased levels of fecal contamination (Kitajima et al. 2018). Several studies consistently indicate that normalizing SARS-CoV-2 concentrations by PMMoV enhances correlations with COVID-19 cases at the community level. For example, Maal-Bared et al. (2023) observed that normalization led to an increase in ρ-values by PMMoV in two wastewater treatment plants (WWTPs). Jafferali et al. (2021) recognized PMMoV as functionally effective, deeming it a potentially suitable internal reference standard for normalizing SARS-CoV-2 concentrations. Zhan et al. (2022) also reported improved correlations with COVID-19 cases when normalizing SARS-CoV-2 levels by PMMoV in campus data. However, an increasing number of reports cast doubt on the assumption that normalization by PMMoV contributes to the improvement of standardization and reporting in wastewater-based epidemiology (WBE) data. Ai et al. (2021) reported that the normalization of the SARS-CoV-2 concentration by PMMoV did not improve the correlation. Feng et al. (2021) reported that SARS-CoV-2 concentrations normalized by PMMoV reduced correlations in 8 out of 12 WWTPs. Nagarkar et al. (2022) also observed that the utility of PMMoV to normalize SARS-CoV-2 concentrations is not universal. These findings highlight the ongoing debate and complexity surrounding using PMMoV as a normalization factor in interpreting SARS-CoV-2 data in WBE. Additionally, although the associations between SARS-CoV-2 RNA concentrations and COVID-19 cases are increasingly reported in the literature, it is noteworthy that most studies primarily concentrate on wastewater treatment plants (WWTPs).

This study aims to assess and compare the effects of PMMoV normalization on the correlations and early warning abilities related to SARS-CoV-2 concentration in raw sewage and various factors, such as active COVID-19 cases, self-isolations, and their combination among all resident students at the University of Tennessee, Knoxville, over the course of one academic year. The findings of this study could enhance our comprehension of the potential applications and limitations of wastewater monitoring for COVID-19 surveillance, particularly within the context of raw sewage, on university campuses.

## Material and Methods

### Raw Sewage Sampling

In the summer of 2020, the University of Tennessee administration made the decision to implement Wastewater-Based Epidemiology (WBE) for the early detection of COVID-19 cases within student residence halls. With each residence hall housing a specific number of students, totaling 7486 individuals in Fall 2020 and 6781 in Spring 2021, these student populations serve as representative indicators reflecting the overall health status of the campus, which served a total of 30,559 students during the 2020 academic year. Raw sewage samples were systematically collected on a weekly basis, encompassing 18 dormitories (housing 6639 individuals in Fall 2020 and 5984 in Spring 2021), 15 fraternities (with 319 occupants in Fall 2020 and 307 in Spring 2021), and 14 sororities (accommodating 528 in Fall 2020 and 490 in Spring 2021), throughout the academic semester from September 14, 2020, to September 21, 2021 (Figure 1, Table 1). The characteristics of the student residence halls are summarized in Table 2-1. However, for the purposes of this study, only the data collected from November 9, 2020, to June 04, 2021, was utilized, corresponding to the period when PMMoV detection started. To ensure targeted sampling from specific buildings with distinct student populations, samples were obtained downstream of dispense valves or sewer manholes just before convergence or mixing with other sewer lines. Grab samples (> 50 ml) were acquired from the manhole using a stainless-steel telescopic rod pole swivel dipper water swing sampler and submerged into the flowing raw sewage, or alternatively, using a sterile Nalgene bottle to collect raw sewage directly from the valve. Sampling started at 8:00 am, and all collected samples were promptly transported to the Biosafety Level-2 (BSL-2) laboratory within a cooler containing ice, ensuring arrival within 3 hours for immediate processing.

**Figure 1.**
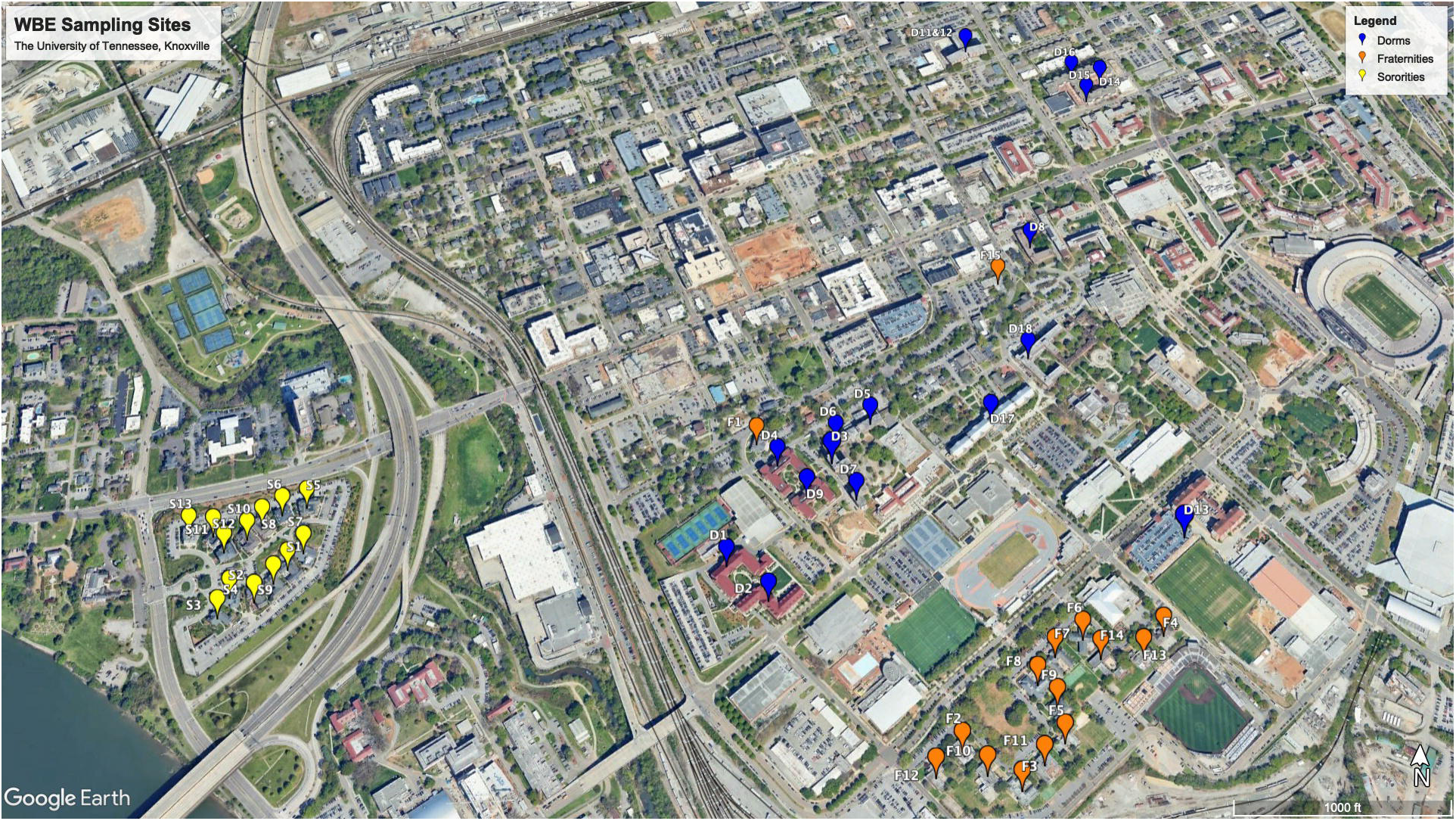
WEB sampling sites in the University of Tennessee, Knoxville.

**Table 1.**
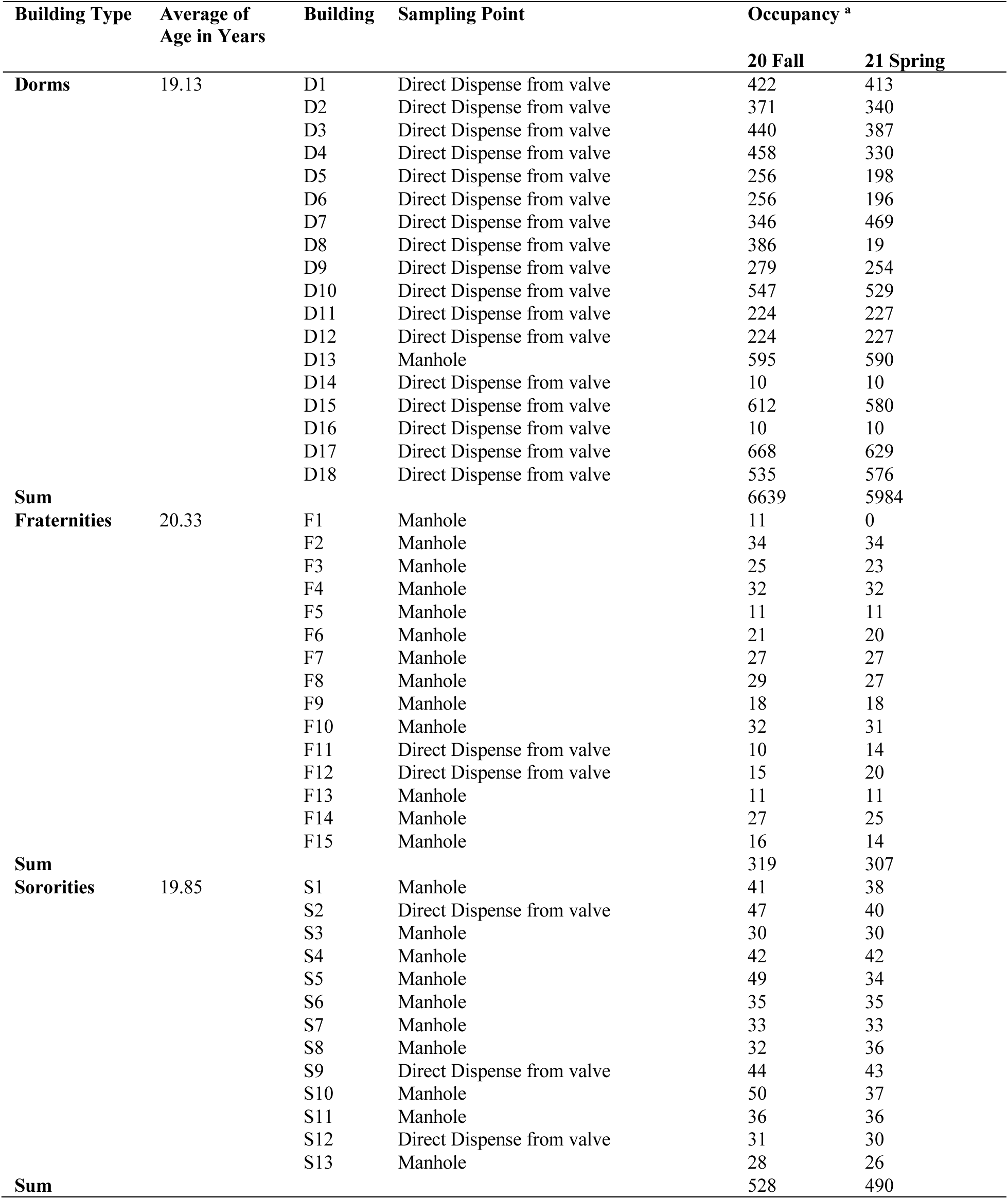
Characteristics of student residence halls in The University of Tennessee, Knoxville. ^a^ Occupancy varied throughout the Fall 2020 and Spring 2021 semester, so the highest numbers of residents at any given time throughout the semester are shown.

**Table 2.**
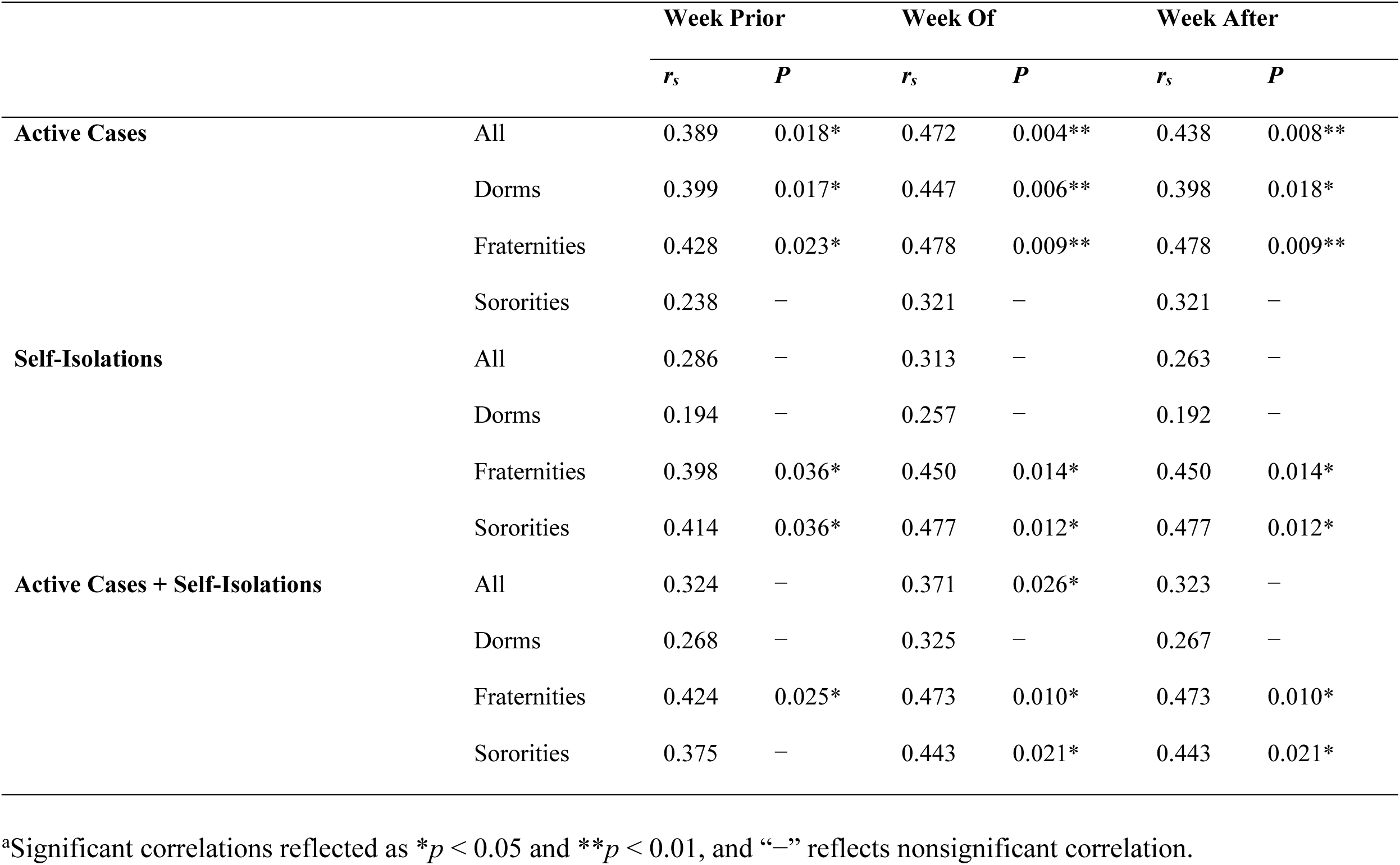
Spearman Correlation Coefficients (*r_s_*) and Significance *p* Values of Wastewater Measures (one-week prior average, at the week, and one-week after) against active, self-isolation and active cases plus self-isolation data without normalization ^a^.

**Table 3.**
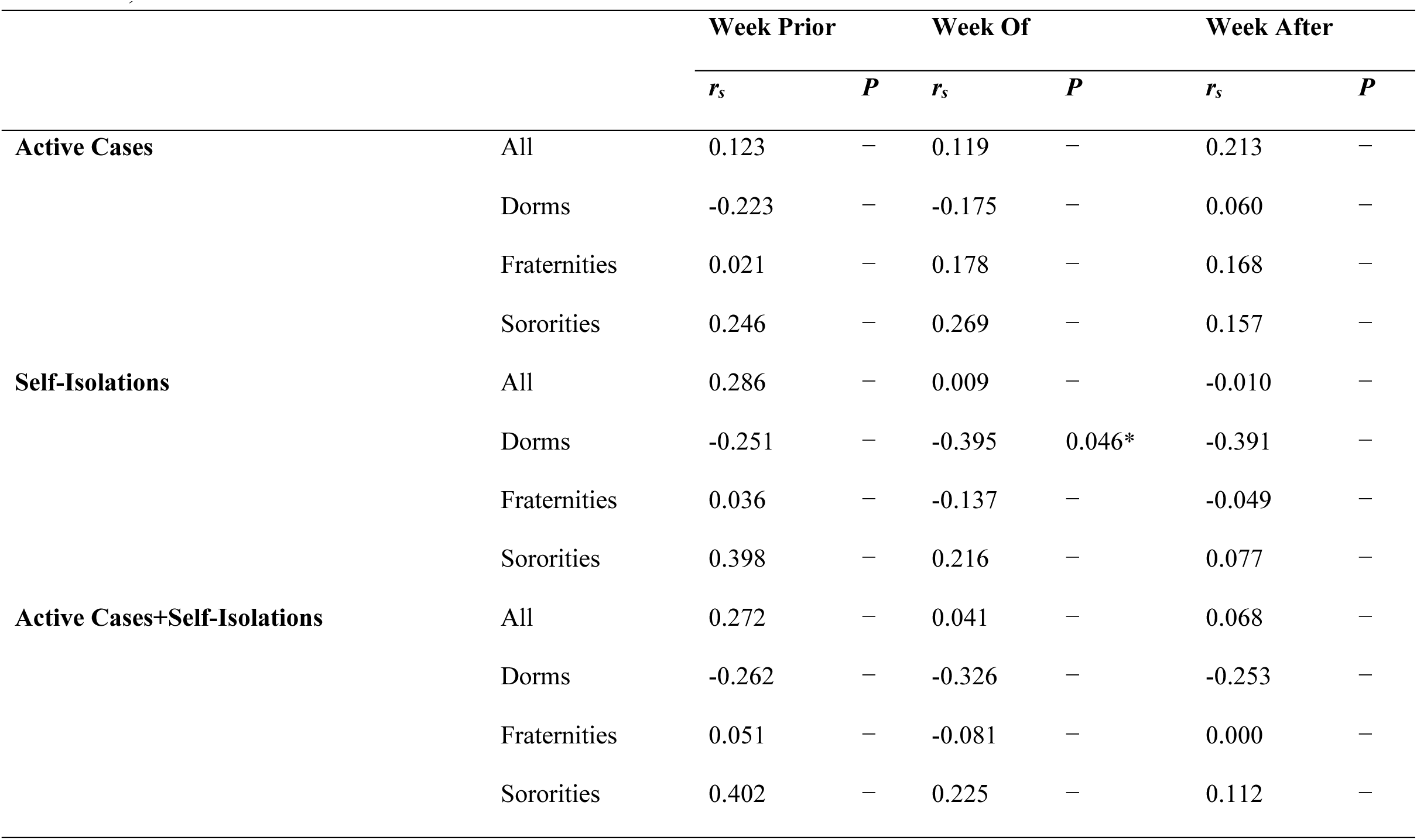
Spearman Correlation Coefficients (*r_s_*) and Significance *p* Values of Wastewater Measures (one week prior average, at the week, and one week after) against active, self-isolation and active cases plus self-isolation data with normalization (SARS-CoV-2/PMMoV) ^a^.

### Sample Processing

The initial concentrations of SARS-CoV-2 and PMMoV RNA were promptly determined from 50 ml of thoroughly mixed raw sewage samples within 5 hours of collection. Subsequently, the raw sewage samples underwent a pasteurization process for 2 hours at 60°C, followed by centrifugation at 5,000 x g for 10 minutes and filtration through 0.45 µm and 0.22 µm nitrocellulose filters to eliminate large suspended particulate matter. The filtered samples were concentrated using ultrafiltration with an Amicon Ultra-15 filtration device (EMD Millipore, Burlington, MA The Amicon Ultra was centrifuged at room temperature at 4,000 x g for 30 minutes (Swing-arm rotor) or 5,000 x g for 20 minutes (Fixed-angle rotor). The resulting concentrate, approximately (∼250 μL), was transferred to 2 mL DNA LoBind tubes, and RNA extractions were performed using a Qiagen viral RNA Mini Kit (Qiagen, Valencia, CA, USA) (Ash et al. 2021). In brief, 60 μL of RNA was extracted from a homogenized sample using the Qiagen viral RNA Mini Kit, following the manufacturer’s instructions. DNase/RNase-free water served as the extraction negative control. All RNA samples were stored at -80°C and subjected to RT-qPCR analysis within one day of RNA extraction.

### RT-qPCR

RT-qPCR was employed to quantify the concentrations of SARS-CoV-2 and PMMoV RNA in each sample. The CDC primer/probe assay for SARS-CoV-2 N1 was utilized, and quantification was conducted using the TaqPath 1-Step RT-qPCR Master Mix, CG (Thermo Fisher Scientific) on an Applied Biosystems QuantStudios 7 Pro Real-Time PCR System instrument (Ash et al. 2023).

Each 20 μL reaction comprised 5 μL of 4X Master Mix (Thermo Fisher Scientific), 0.25 μL of 10 μmol/L probe, 1 μL each of 10 μmol/L forward and reverse primers, 7.75 μL of nuclease-free water, and 5 μL of nucleic acid extract. The reagents were pipetted into 96-well plates and vortexed for 10 seconds. Thermocycling conditions were as follows: uracil-DNA glycosylase incubation for 2 minutes at 25°C, reverse transcription for 15 minutes at 50°C, activation of the Taq enzyme for 2 minutes at 95°C, and two-step cycling for 3 seconds at 95°C and 30 seconds at 55°C for 45 cycles. A positive test result was defined as an exponential fluorescent curve that crossed the threshold within 40 cycles (cycle threshold [Ct] <40).

PMMoV quantification was performed using RT-qPCR with the TaqPath 1-Step RT-qPCR Master Mix, CG (Thermo Fisher Scientific) on a QuantStudios 7 Pro instrument. In each 20 μL reaction, the components included 5 μL of 4X Master Mix (Thermo Fisher Scientific), 0.5 μL of 10 μmol/L probe, 1.8 μL each of 10 μmol/L forward and reverse primers, 8.9 μL of nuclease-free water, and 2 μL of nucleic acid extract. The reagents were pipetted into 96-well plates and vortexed for 10 seconds. Thermocycling conditions were as follows: uracil-DNA glycosylase incubation for 2 minutes at 25°C, reverse transcription for 15 minutes at 50°C, activation of the Taq enzyme for 10 minutes at 95°C, and two-step cycling for 30 seconds at 95°C and 1 minute at 60°C for 40 cycles.

Each RT-qPCR run incorporated a series of three positive and negative controls, where the positive control comprised Mastermix and DNase/RNase-free water. All RT-qPCR reactions were conducted in triplicate, and results were considered valid only if the positive control yielded a positive outcome and the negative control remained negative. A sample was deemed positive only when all replicates were positive, each falling within the linear range of the standard curve.

The efficiency of the N1 standard curve was determined to be 94.7% (R^2^ = 1). The final quantification of SARS-CoV-2 RNA was calculated as the mean of three replicates of virus copies. The RT-qPCR outputs were subsequently converted to copies per liter. Notably, the detection limit for SARS-CoV-2 and PMMoV in this study was established at 20 and 10 copies/L, respectively (Bustin et al. 2009).

### Data Analysis

Statistical analysis was conducted to investigate the relationship between SARS-CoV-2 concentrations and reported cases during a significant event. Data reflecting daily new cases at residence-specific levels were utilized for statistical analysis from November 9, 2020, to June 04, 2021. To facilitate the analysis, weekly case counts were aggregated in three ways: raw sewage sample collection one week before, during the week of, and one week after active cases and self-isolations. Spearman’s correlation assessed the relationship between raw and PMMoV-normalized SARS-CoV-2 concentrations and the aggregated case counts. The use of Spearman’s correlation avoided the assumptions of normality and the absence of outliers associated with the Pearson correlation. The correlation coefficient’s magnitude indicated the association’s strength and direction, measuring how closely the points aligned along the monotonic association. Non-detects in viral data were treated as twenty values for statistical purposes.

## Results

### Active and isolation clinical COVID-19 cases

The Student Health Center provides valuable insights into the prevalence of COVID-19 on campus, reporting both active and isolated cases. Throughout the study period, an analysis of the data reveals a cumulative total of 2,321 active cases and 4,202 isolated cases spread across the 46 residence halls from September 14, 2020, to June 04, 2021 (Figure 3). The onset of the pandemic at the campus was marked by a significant surge in COVID-19 cases during the initial wave in September, with 778 active cases and 1,480 isolated cases. An in-depth analysis of specific timeframes throughout the study period reveals distinct waves of COVID-19 incidence. Following the September surge, subsequent months displayed varying degrees of activity. In October, there were 208 active cases and 345 isolation cases, indicating a notable decline from the initial peak. November saw a resurgence with 310 active cases and 628 isolation cases, suggesting a renewed increase in viral spread. December recorded 144 active cases and 55 isolation cases, signaling a potential decrease in cases toward the end of the year. As the new year began, January experienced a further rise with 170 active cases and 161 isolation cases. The trend continued in February with 292 active cases and 663 isolation cases, reaching another peak. March and April demonstrated a gradual decline, with 288 active cases, 757 isolation cases, 125 active cases, and 185 isolation cases, respectively. May saw a minimal presence, with only five active cases and two isolation cases, indicating a substantial reduction in COVID-19 incidence. Notably, during the winter break, spanning from December 12, 2020, to January 13, 2021, there remained a cumulative total of 160 active cases and 71 isolation cases, underscoring the persistent impact of the virus even during the holiday period.

### The concentration of SARS-CoV-2 and PMMoV at different building types

The university implemented raw wastewater surveillance as a complementary strategy to clinical testing, aiming to detect the presence of SARS-CoV-2 on campus. This approach is particularly valuable in capturing individuals who may be asymptomatic or have mild symptoms and thus might not actively seek clinical screening. A comprehensive overview of the Wastewater-Based Epidemiology (WBE) project at the University of Tennessee, Knoxville, is provided in our paper by Ash et al. (2023). Figures 3, 4, 5, and 6 illustrate the measured viral copies per liter (copies/L) of the N1 genes across all residence halls, dormitories, fraternities, and sororities. The concentrations of SARS-CoV-2 RNA in raw sewage samples were 2.75 × 10^2^ ± 5.49 × 10^2^ copies/L for all residence halls, 1.97 × 10^2^ ± 3.48 × 10^2^ copies/L for dormitories, 6.93 × 10^2^ ± 2.12 × 10^3^ copies/L for fraternities, and 1.30 × 10^2^ ± 1.91 × 10^2^ copies/L for sororities. Correspondingly, the concentrations of PMMoV RNA across all samples were 1.55 × 10^4^ ± 2.79 × 10^4^, 1.57 × 10^4^ ± 1.20 × 10^4^, 3.07 × 10^4^ ± 9.98 × 10^4^, and 5.26 × 10^3^ ± 2.74 × 10^3^ copies /L for all residence halls, dormitories, fraternities, and sororities, respectively.

### All residence halls

Analyzing data of all residence halls (covering 18 dormitories, 15 fraternities, and 14 sororities) from November 2020 to May 2021, the temporal dynamics revealed fluctuating concentrations (Figure 3). Notably, in November 2020, the concentration was 3.06 × 10^2^ ± 1.70 × 10^2^ copies/L, followed by a decrease in December 2020 to 2.74 × 10^1^ ± 1.28 × 10^1^ copies/L. January 2021 showed a slight increase to 5.74 × 10^1^ ± 2.90 × 10^1^ copies/L, while February 2021 witnessed a notable rise to 7.61 × 10^2^ ± 1.27 × 10^3^ copies/L. Subsequent months displayed varying concentrations, with March at 3.28 × 10^2^ ± 4.21× 10^2^ copies/L, April at 2.43 × 10^2^ ± 3.55 × 10^2^ copies/L, and May at non-detectable.

The concentration of PMMoV also exhibited variations across all residence halls during the same period (Figure 2-3). In November 2020, the PMMoV concentration was 9.84 × 10^3^ ± 4.32 × 10^3^ copies/L, increasing in December 2020 to 1.08 × 10^4^ ± 1.28 × 10^4^ copies/L. January 2021 recorded a further increase to 1.59 × 10^4^ ± 1.70 × 10^3^ copies/L. February 2021 exhibited a relatively stable concentration at 1.50 × 10^4^ ± 6.53 × 10^3^ copies/L. March 2021 substantially increased to 4.15 × 10^4^ ± 5.97 × 104 copies/L. April 2021 decreased to 5.04 × 10^3^ ± 2.53 × 10^3^ copies/L, and in May 2021, the concentration was 2.63 × 10^3^ ± 4.97 × 10^3^ copies/L.

**Figure 2.**
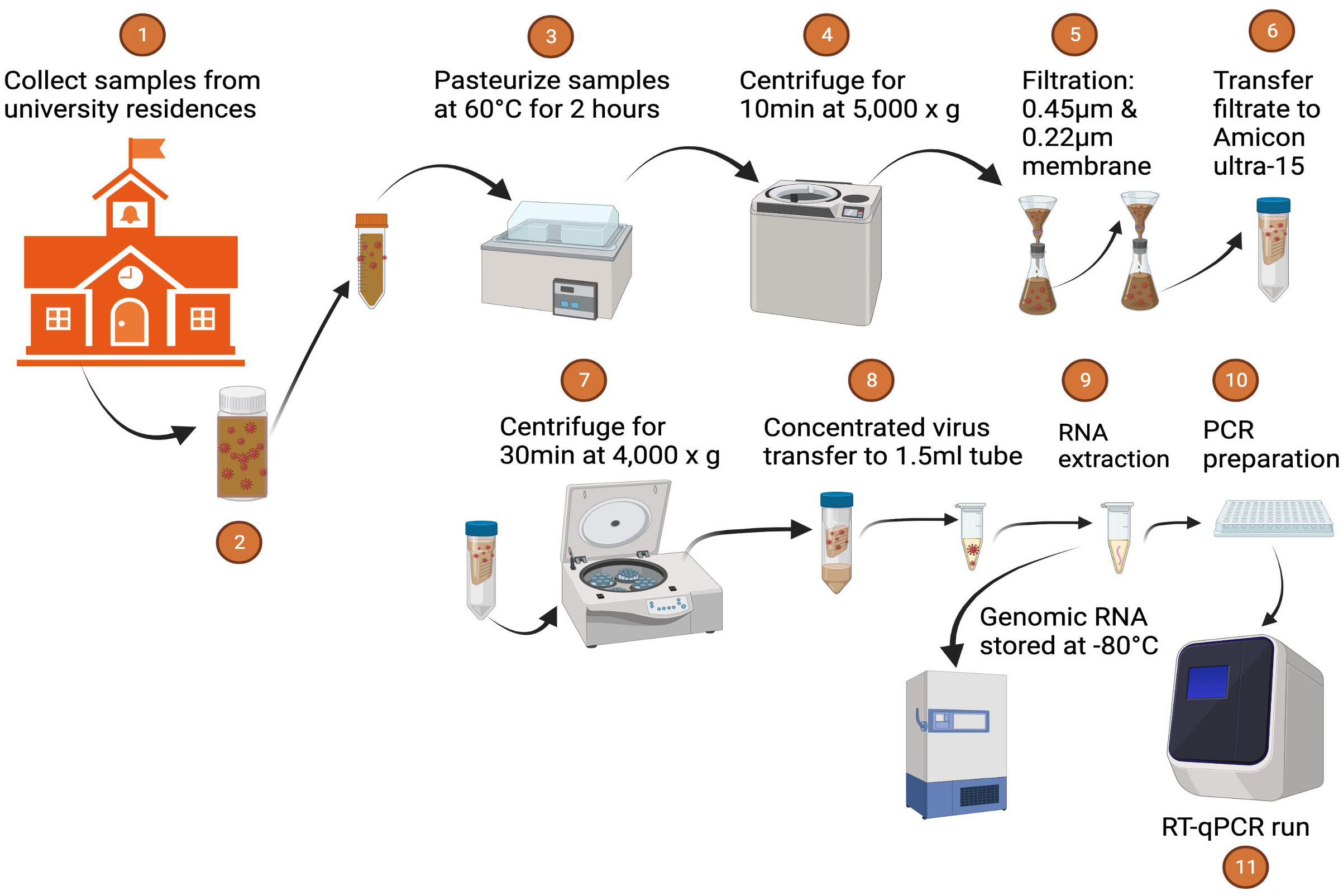
Sampling and Processing.

**Figure 3.**
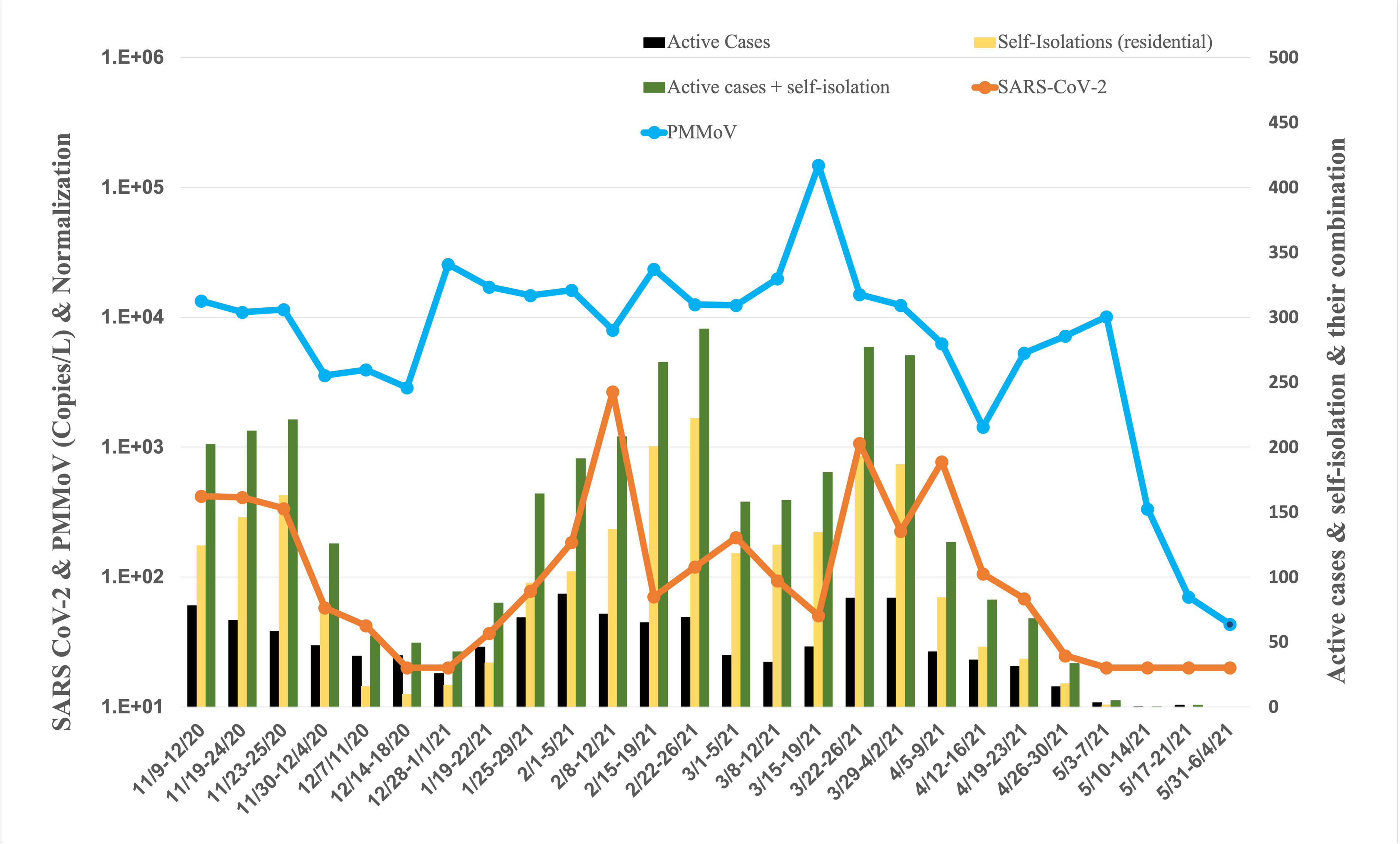
Average Concentration of SARS-CoV-2 in all residence halls, and active cases, and self-isolation students in the residential halls.

### Dormitories

The concentration of SARS-CoV-2 in 18 dormitories exhibited temporal variations from November 2020 to May 2021 (Figure 4). In November 2020, the concentrations were 2.98 × 10^2^ ± 1.56 × 10^2^ copies per liter (copies/L), indicating a moderate level. This was followed by a notable decrease in December 2020, with a concentration of 2.74 × 10^1^ ± 1.28 × 10^1^ copies/L, suggesting a significant reduction. January 2021 showed a slight increase to 7.16 × 10^1^ ± 9.22 × 10^1^ copies/L, followed by a moderate rise in February 2021, with a concentration of 1.94 × 10^2^ ± 1.65 × 10^3^ copies/L. March 2021 witnessed a similar level at 1.49 × 10^2^ ± 1.66× 10^2^ copies/L, while April 2021 displayed a substantial increase to 5.25 × 10^2^ ± 8.23 × 10^2^ copies/L. In May 2021, the concentration decreased to a non-detectable level.

**Figure 4.**
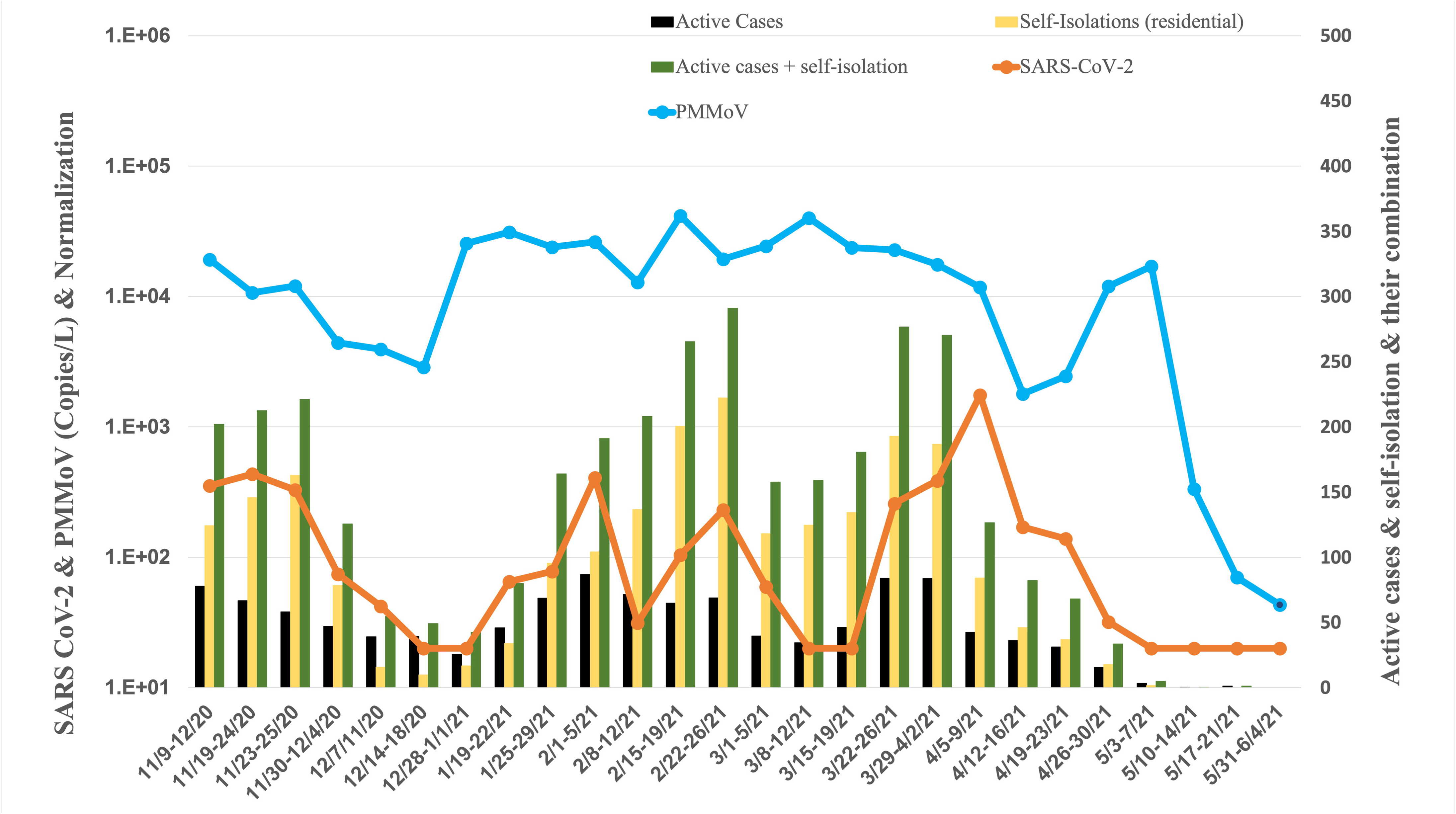
Average Concentration of SARS-CoV-2 in dorms, and active cases, and self-isolation student in the residential halls.

Concurrently, the concentration of PMMoV in dormitories exhibited variations from November 2020 to May 2021 (Figure 4). In November 2020, the PMMoV concentration was 1.16 × 10^4^ ± 6.08 × 10^3^ copies/L. December 2020 showed a similar level at 1.08 × 10^4^ ± 1.28 × 10^4^ copies/L. January 2021 increased to 2.75 × 10^4^ ± 5.10 × 10^3^ copies/L. February 2021 exhibited a relatively stable concentration at 2.50 × 10^4^ ± 1.23 × 10^4^ copies/L. March 2021 displayed a similar level at 2.57 × 10^4^ ± 8.42 × 10^3^ copies/L. April 2021 decreased to 7.00 × 10^3^ ± 5.63 × 10^3^ copies/L, and in May 2021, the concentration was 4.38 × 10^3^ ± 8.45 × 10^3^ copies/L.

### Fraternities

The concentration of SARS-CoV-2 in 15 fraternities exhibited dynamic changes from November 2020 to May 2021 (Figure 5). In November 2020, the concentration was 1.80 × 10^2^ ± 2.53× 10^2^ copies per liter (copies/L). January 2021 decreased to 2.36 × 10^1^ ± 1.89 × 10^0^ copies/L, indicating a substantial reduction. February 2021 recorded a significant increase to 2.34× 10^3^ ± 4.62 × 10^3^ copies/L, suggesting a resurgence. The concentration remained elevated in March 2021 at 6.60 × 10^2^ ± 1.30 × 10^3^ copies/L. April 2021 displayed a moderate level at 11.06 × 10^2^ ± 1.21× 10^2^ copies/L; in May 2021, the concentration was non-detectable.

**Figure 5.**
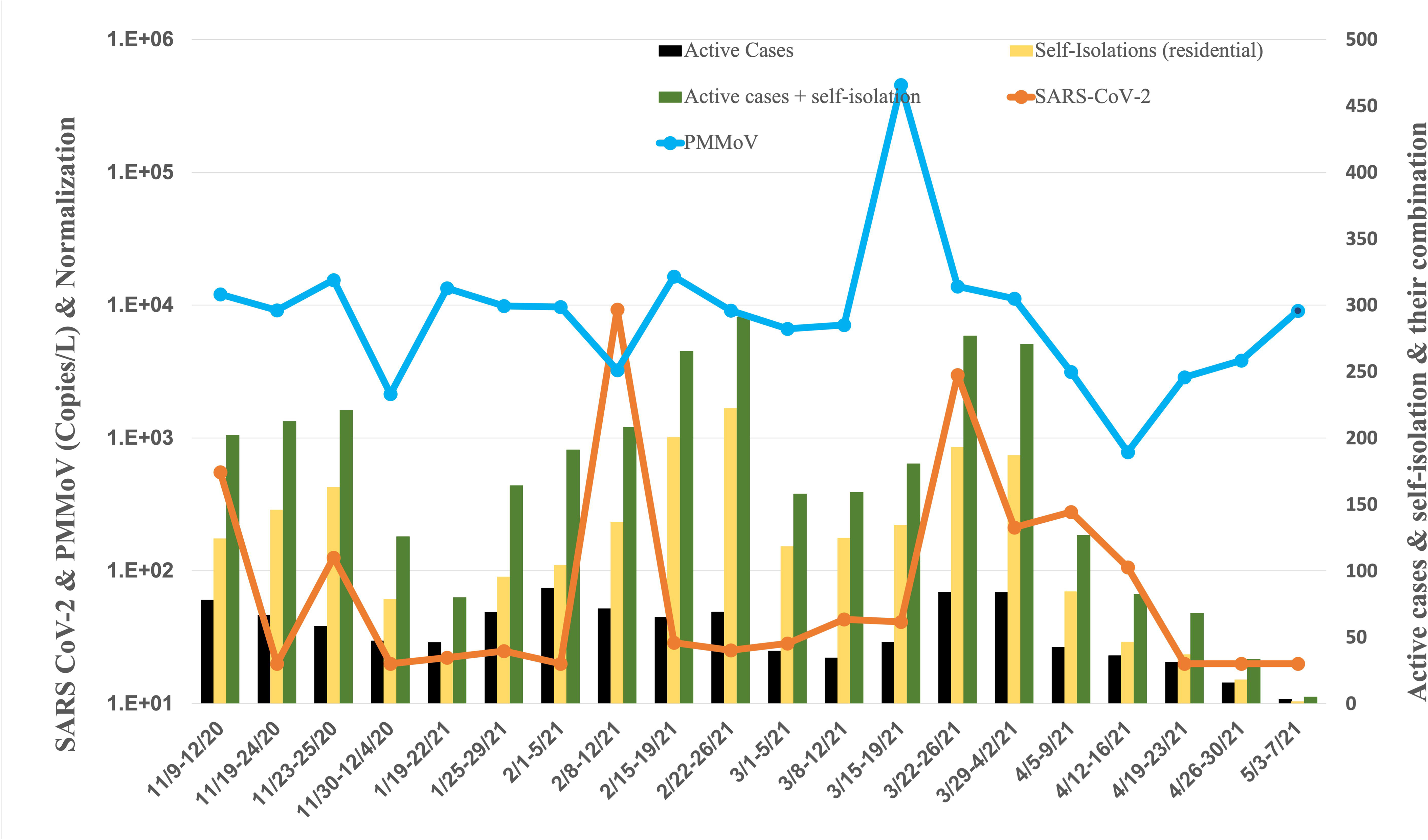
Average Concentration of SARS-CoV-2 in fraternity, and active cases, and self-isolation student in the residential halls.

Simultaneously, the concentration of PMMoV in fraternities exhibited varying levels from November 2020 to May 2021 (Figure 2-5). In November 2020, the PMMoV concentration was 9.71 × 10^3^ ± 5.67 × 10^3^ copies/L. January 2021 increased to 1.16 × 10^4^ ± 2.50 × 10^3^ copies/L. February 2021 exhibited a similar level at 9.62 × 10^3^ ± 5.40 × 10^3^ copies/L. March 2021 recorded a substantial increase to 9.86 × 10^4^ ± 1.99 × 10^5^ copies/L. April 2021 displayed a decrease to 2.66 × 10^3^ ± 1.31 × 10^3^ copies/L. In May 2021, the concentration was 9.07 × 10^3^ copies/L.

### Sororities

The concentration of SARS-CoV-2 in 14 sororities displayed varying levels from November 2020 to May 2021 (Figure 6). In November 2020, the concentration was 3.46 × 10^2^ ± 3.18× 10^2^ copies per liter (copies/L). January 2021 decreased to 7.34 × 10^1^ ± 7.55 × 10^1^ copies/L, indicating a substantial reduction. February 2021 recorded a further decrease to 4.41× 10^1^ ± 2.78× 10^1^ copies/L. March 2021 exhibited a modest increase to 2.02 × 10^2^ ± 2.15× 10^2^ copies/L. April 2021 displayed a lower concentration at 2.09 × 10^1^ ± 1.79× 10^0^ copies/L. In May 2021, the concentration was non-detectable, signifying a significant decrease or absence of viral presence in the raw sewage samples from sororities during that specific month.

**Figure 6.**
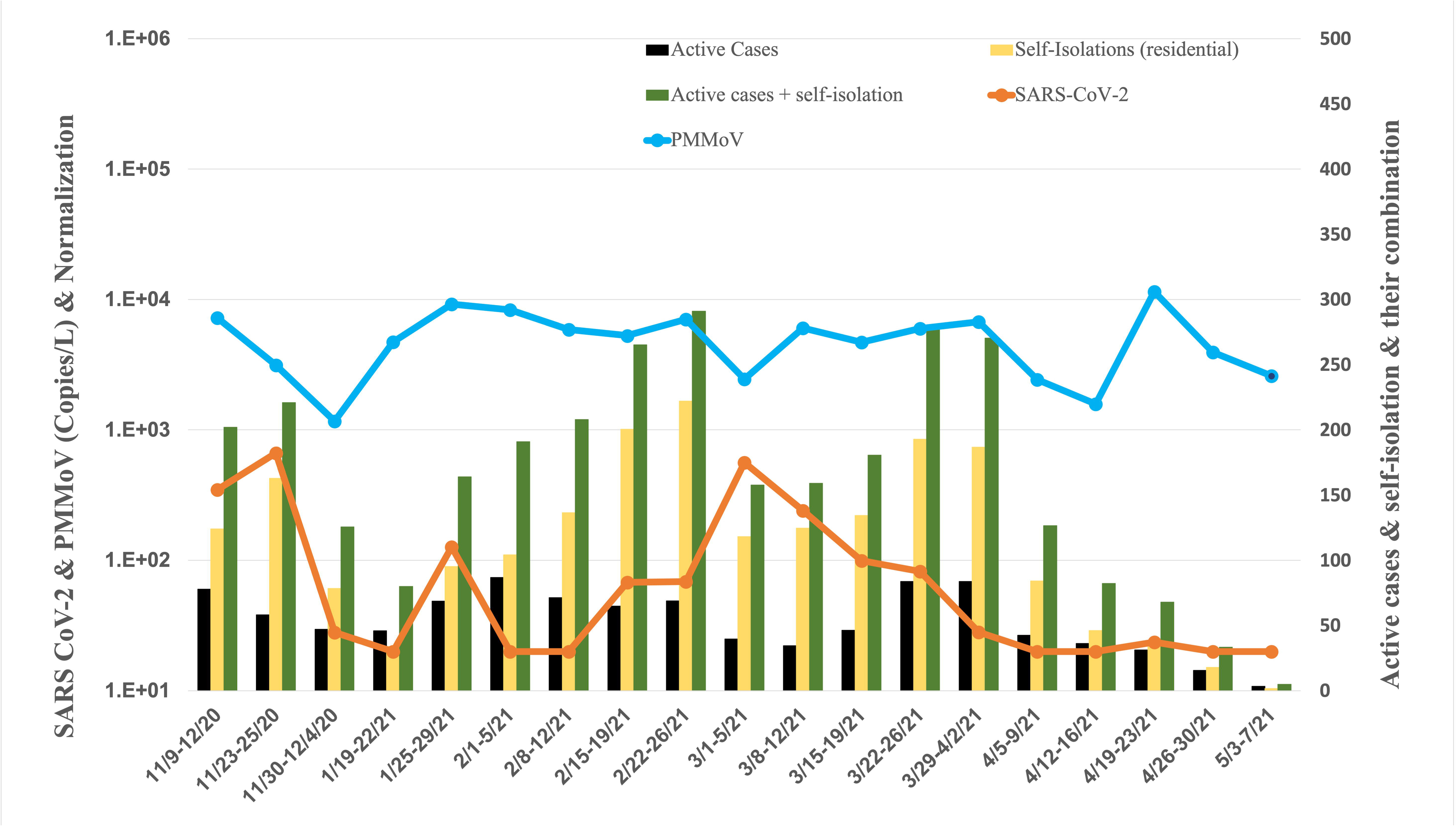
Average Concentration of SARS-CoV-2 in sororities, active cases, and self-isolation students in the residential halls

Concurrently, the concentration of PMMoV in sororities displayed varying levels from November 2020 to May 2021 (Figure 6). In November 2020, the PMMoV concentration was 3.84 × 10^3^ ± 3.09 × 10^3^ copies/L. January 2021 increased to 6.97 × 10^3^ ± 3.81 × 10^3^ copies/L. February 2021 exhibited a similar level at 6.64 × 10^3^ ± 1.35 × 10^3^ copies/L. March 2021 decreased to 5.18 × 10^3^ ± 1.70 × 10^3^ copies/L. April 2021 displayed a further decrease to 4.86 × 10^3^ ± 4.53 × 10^3^ copies/L. In May 2021, the concentration was 2.60 × 10^3^ copies/L.

### Impacts of normalization on correlations between SARS-CoV-2 concentration and COVID-19 cases

Tables 1 and 2 provide a comprehensive summary of the Spearman correlation (*r_s_*) values, elucidating the strength of associations between unnormalized and normalized SARS-CoV-2 concentrations by PMMoV and various factors such as active cases, self-isolations, and the combination of active cases with self-isolations. The analysis encompasses all residence halls, dormitories, fraternities, and sororities. Interpretation of Spearman’s ρ values adheres to the criteria outlined by von Sperling et al. (2020). Correlations with a ρ-value exceeding 0.7 are considered strong; those ranging between 0.4 and 0.7 indicate moderate correlations and values falling between 0 and 0.4 denote weak associations. Negative values signify weakened associations, while positive values suggest improved associations.

Active cases, both independently and in combination with self-isolations, demonstrated noteworthy correlations with SARS-CoV-2 concentrations across all residential halls, spanning from moderate to weak levels. Post-normalization, the correlations involving either active cases alone or the combined metric of active cases with self-isolations and SARS-CoV-2 concentrations diminished, revealing no statistically significant associations.

In dormitories, active cases displayed significant correlations with SARS-CoV-2 concentrations, characterized by moderate levels. Nevertheless, these correlations waned after normalization by PMMoV, resulting in statistically insignificant associations with SARS-CoV-2 concentrations.

Within fraternities, active cases, self-isolations, and their combination exhibited substantial correlations with SARS-CoV-2 concentrations, marked by moderate correlation levels. However, normalization by PMMoV led to a reduction in these correlations, rendering the associations with SARS-CoV-2 concentrations statistically insignificant.

Similarly, self-isolations and the combined metric of active cases with self-isolations showed significant correlations with SARS-CoV-2 concentrations within sororities, characterized by moderate correlation levels. Yet, normalization by PMMoV resulted in a reduction of these correlations, leading to statistically insignificant associations with SARS-CoV-2 concentrations.

### Impacts of normalization correlations and early warning of COVID-19 cases trend

Tables 1 and 2 elucidate the differences in *r_s_* values between unnormalized and normalized datasets. These values encapsulate the correlations between SARS-CoV-2 concentrations and various factors including active cases, self-isolations, and the combination of active cases with self-isolations. The analysis includes both a seven-day advance and a seven-day lag to discern lead times between WBE data and clinical cases, specifically emphasizing diverse student residence halls.

Preceding SARS-CoV-2 concentrations by one week demonstrated weakly significant correlations with active cases across all residential halls, while one week after, moderate correlations were observed with active cases. Importantly, weakly insignificant correlations were noted between the one-week advance and one-week lag of SARS-CoV-2 concentrations and both self-isolations and the combination with active cases.

Similarly, a seven-day advance and a seven-day lag of SARS-CoV-2 concentrations revealed weakly significant correlations with active cases across dormitories. Notably, weakly insignificant correlations were observed between the one-week advance and one-week lag of SARS-CoV-2 concentrations and both self-isolations and the combination with active cases.

A seven-day advance and a seven-day lag of SARS-CoV-2 concentrations exhibited moderately significant correlations with active cases and the combination with self-isolations among fraternities. Furthermore, weakly significant correlations were identified between the one-week advance of SARS-CoV-2 concentrations and self-isolations, while a moderate correlation was observed for the one-week lag.

A seven-day advance and a seven-day lag of SARS-CoV-2 concentrations exhibited moderately significant correlations with self-isolations among sororities. Furthermore, moderate significant correlations were identified between the one-week lag of SARS-CoV-2 concentrations and the combination of active cases and self-isolations.

Normalization by PMMoV resulted in either a reduction or an insignificant impact on *r_s_* values for a seven-day advance and a seven-day lag of SARS-CoV-2 concentrations with active cases, self-isolations, and their combinations across all building types in this study.

## Discussion

### SARS-CoV-2 concentrations in raw wastewater reflect the trends of COVID-19

Integrating wastewater-based epidemiology (WBE) into public health surveillance is progressing as a strategic response to the challenges posed by the COVID-19 pandemic. A crucial aspect of this approach is understanding the correlations between SARS-CoV-2 concentrations and COVID-19 cases, particularly in communities characterized by mobile populations and significant temporal fluctuations. This significance is accentuated in situations where reported clinical case numbers may not accurately reflect the actual residents in these communities.

In our study, we observed notable variations in SARS-CoV-2 concentrations among different building types, irrespective of whether they accommodated high (>200) or low (<50) student numbers in the corresponding residence halls. Our findings underscore a consistent alignment between SARS-CoV-2 concentrations in raw sewage from all student residence halls, as assessed through RT-qPCR, and the trends observed in active cases of COVID-19 across all residence halls. These results are in harmony with existing research, affirming that SARS-CoV-2 concentrations serve as an effective tool for estimating the dynamics of COVID-19 cases within diverse communities. Medema et al. (2020) reported a significant correlation between SARS-CoV-2 concentration and cumulative COVID-19 prevalence, even in situations of low prevalence in the Netherlands. Xiao et al. (2022) documented that SARS-CoV-2 concentration preceded new cases during the first wave of COVID-19 in MA, USA. Additionally, Ai et al. (2021) found a robust correlation between SARS-CoV-2 concentration and confirmed cases in 8 sewersheds. Wurtzer et al. (2020) similarly reported correlations between SARS-CoV-2 concentrations and the number of symptomatic or non-symptomatic COVID-19 patients at local or regional scales. Furthermore, Schmitz et al. (2021) discovered a correlation between clinical data and corresponding sewage SARS-CoV-2 concentrations in student dorms at the University of Arizona during the fall semester of 2020.

Given that dormitories, fraternities, and sororities represented as microcosms of the campus, our study revealed that the SARS-CoV-2 concentrations identified in dormitories and fraternities correspondingly mirrored the trends observed in COVID-19 cases across all student residence halls. The greater proportion of undergraduates residing in dormitories might provide a more complete picture of the COVID-19 situation on campus. Fraternities may be factored into the analysis due to the correlation between males and the presence of fecal viruses (Daou et al. 2022). This correlation is consistent with the research conducted by Bitter et al. (2022), wherein the university was identified as a microcosm of the corresponding city and exhibited analogous trends in COVID-19 cases.

Furthermore, our analysis revealed that Spearman rank correlations ranged from ρ = 0.447 to 0.478, with no relation to the size of the student residence halls, consistent with the findings of Feng et al. (2021), who noted similar correlations unrelated to the size of wastewater treatment plants.

### Normalization by PMMoV did not strengthen the correlation between SARS-CoV-2 concentration and COVID-19 trend

While the normalization of SARS-CoV-2 concentrations in wastewater by PMMoV has the potential to enhance comparability across studies, our investigation demonstrated that there was no substantial improvement in correlations between SARS-CoV-2 concentrations in raw wastewater and active cases, self-isolation, or their combination when compared to unnormalized data, irrespective of the student population in the respective residence halls. The results of this study add to the expanding body of research that questions the notion that normalization by PMMoV enhances the reporting and standardization of WBE data. Ai et al. (2021) reported that normalization by PMMoV did not significantly improve correlations with new case numbers or enhance estimation models across nine wastewater treatment plants serving populations ranging from 14,000 to 900,000. Similarly, Feng et al. (2021) found that normalizing SARS-CoV-2 concentrations to PMMoV did not improve correlation coefficients in all 12 WWTPs serving populations from 11,000 to 616,000. In the study by Greenwald et al. (2021), normalization by PMMoV led to a reduction in significant correlations to clinical cases in wastewater catchment areas serving populations from 2,930 to 1,500,000. A recent study by Nagarkar et al. (2022) found that PMMoV had a stronger correlation with samples from a larger sewershed, which served approximately 488,000 people and had higher levels of industrial and stormwater inputs. Zambrana et al. (2022) found that normalizing SARS-CoV-2 concentrations by PMMoV did not substantially change the correlation coefficients at various levels, including campus level (serving around 10,000 people), building level (serving around 500 people), or individual-building level (serving around 200 people) within the Stanford University campus. These findings contrast with other studies suggesting that normalization to PMMoV enhances relationships with clinical data. It is crucial to note that these divergent conclusions were primarily attributed to the choice of matrix (solids versus liquid) (D’Aoust et al. 2021, Wolfe et al. 2021) and variances in analytical methods (Scott et al. 2021, Zhan et al. 2022).

Environmental uncertainties introduce complexities in normalizing SARS-CoV-2 concentrations in raw wastewater by PMMoV. Raw wastewater from university dormitories primarily consists of washing and bathing effluents, occasionally mixed with kitchen wastewater. In contrast, sewage wastewater derives from a broader spectrum, including residential sources (private residences, dormitories, hotels, and residential care facilities) and commercial facilities (including hospitals), creating a more intricate composition compared to university dormitory wastewater. Hamouda et al. (2021) underscored the role of microorganisms and physico-chemical properties, such as pH, solids, and disinfectants, in influencing the persistence and detection of SARS-CoV-2 RNA in wastewater. These factors impact the genetic material’s integrity, posing challenges in detection. Higher pH in wastewater has been linked to decreased concentrations of all three SARS-CoV-2 targets (Ahmed et al. 2020, Maal-Bared et al. 2023). Sapula et al. (2021) reported the non-detection of SARS-CoV-2 in a wastewater plant with elevated pH levels (8.80 to 9.35). These findings also have implications for wastewater treatment plants receiving waste with high pH (pH > 7.75) from various sources, including lagoons, septic tanks, and industrial operations. Elevated pH levels can significantly affect virus adsorption to particles and the concentrations recovered, emphasizing the need for careful consideration of wastewater composition and pH variations in interpreting surveillance data. Accurate interpretation of WBE relies on addressing environmental uncertainties in both raw wastewater and influent wastewater.

The correlation between SARS-CoV-2 concentrations and PMMoV normalization may be influenced by the distinct characteristics of the two viruses in terms of their source and origin. SARS-CoV-2 is an enveloped virus, while PMMoV is non-enveloped. In contrast to wastewater transported to treatment plants, our study collected wastewater directly from manholes next to the buildings or from access points before the sewer pipe left the building and promptly processed it in the lab. This immediate processing could affect virus concentrations due to variations in decay rates and variability between the enveloped SARS-CoV-2 and non-enveloped PMMoV. Research conducted by Li et al. (2023) has indicated a faster decay rate of SARS-CoV-2 RNA compared to PMMoV RNA in raw sewage. PMMoV RNA exhibited higher abundance and lower variability than pathogenic SARS-CoV-2 RNA (Li et al. 2023). A study by Ye et al. (2016) found that up to 26% of enveloped viruses (murine hepatitis virus and *Pseudomonas* phage φ6) adsorbed to the solid fraction of wastewater, compared to 6% of non-enveloped viruses (Enterobacteria phage MS2 and T3). This implies that enveloped viruses may maintain greater integrity in wastewater with higher total suspended solids (TSS) and turbidity levels. Recognizing the distinct characteristics of enveloped and non-enveloped viruses is crucial for enhancing the precision and meaningfulness of interpretation the data in the context of WBE studies.

### Early warning without and with normalization by PMMoV

The study investigated the impact of normalizing SARS-CoV-2 concentrations by PMMoV on lead times associated with active cases, self-isolation, or their combination. The significant correlations observed one week prior when analyzing SARS-CoV-2 concentrations from all residence halls in this study, indicating a potential early warning utility. These results suggest that SARS-CoV-2 concentrations may serve as an early indicator, aligning with findings from other studies. Nemudryi et al. (2020) reported that SARS-CoV-2 concentration in wastewater precedes clinical test results by 2 to 4 days at the community level. Greenwald et al. (2021) found that the strongest correlation between clinical testing data and SARS-CoV-2 concentration was associated with a two-week lead time across six wastewater catchment areas. Karthikeyan et al. (2021) observed a strong correlation between SARS-CoV-2 concentration in raw untreated wastewater and clinically cases at the county level. The integration of this correlation, along with historical reported case numbers and temporal information in an autoregressive integrated moving average model, facilitated the successful prediction of new reported cases up to 3 weeks in advance. Wu et al. (2022) found that SARS-CoV-2 RNA concentrations in wastewater were correlated with clinically diagnosed new COVID-19 cases, revealing trends four to ten days earlier in wastewater than in clinical data during the initial wave of the pandemic at the Deer Island Wastewater Treatment Plant. However, Xiao et al. (2022) observed that this leading indicator effect was not apparent in the second wave from the same treatment plant. The authors attributed this change to the expansion of testing capacity, enabling more timely identification, and reporting of cases. Additionally, the diverse approaches to reporting cases, considering factors like disease onset, test date, and test result date, also contribute to the variability observed in lead times. Ai et al. (2021) emphasized this by noting that their clinical cases staggered the trend of SARS-CoV-2 concentrations by 3 days since positive cases were assigned a date based on the estimate of disease onset.

In the present study, normalizing SARS-CoV-2 concentrations by PMMoV did not result in enhancements of lead times associated with active cases, self-isolation, or their combination across all examined locations. This observation aligns with the findings of Maal-Bared et al. (2023), who reported that normalizing SARS-CoV-2 concentrations by PMMoV not improved lead times with active cases in ten out of twelve communities.

The significant correlations observed one week prior of SARS-CoV-2 concentrations with active cases, self-isolation, or their combination in all student residence halls in this study but found no improvement in the correlations. The strongest correlations were observed for comparisons with SARS-CoV-2 concentrations during the week of and the week after the measurement period. Consistent with our findings, Ai et al. (2021) also reported no enhancement in correlations for the estimation of leading times. This lack of improvement may be attributed to viral load variations associated with disease progression (Benefield et al. 2020, Walsh et al. 2020). Zheng et al. (2020) further emphasized that stool sample viral loads were highest during the third and fourth weeks after disease onset.

## Conclusion

This year-long study monitored SARS-CoV-2 concentrations in the raw sewage of a university campus, aiming to understand the correlations with various aspects, including active COVID-19 cases, self-isolations, and their combination among all residence students. The findings revealed significant positive correlations between SARS-CoV-2 concentrations a week prior and during the week of monitoring from all student residence halls, dormitories, and fraternities and the occurrence of active COVID-19 cases among all residence students. This suggests that raw sewage monitoring can be a valuable tool for early warning and outbreak tracking within the campus setting. However, normalizing data using the fecal marker PMMoV did not demonstrate clear utility in interpreting campus-wide data. These results contribute to our understanding of the potential applications and limitations of wastewater monitoring for COVID-19 surveillance within the context of raw sewage on university campuses.

## Data Availability

All data produced in the present study are available upon reasonable request to the authors

## Acknowledgments

We extend our gratitude to Mike Humphrys from the Institute for Genome Sciences, School of Medicine, University of Maryland-Baltimore, for providing valuable insights into the selection of RT-qPCR assays and genome sequencing methods. Furthermore, we appreciate the guidance received from Betancourt at the University of Arizona. The funding for this project was generously provided by the University of Tennessee Office of Research and Engagement.

## References

Ahmed, W., Angel, N., Edson, J., Bibby, K., Bivins, A., O’Brien, J. W., Choi, P. M., Kitajima, M., Simpson, S. L., Li, J., Tscharke, B., Verhagen, R., Smith, W. J. M., Zaugg, J., Dierens, L., Hugenholtz, P., Thomas, K. V. and Mueller, J. F. (2020). “First confirmed detection of SARS-CoV-2 in untreated wastewater in Australia: A proof of concept for the wastewater surveillance of COVID-19 in the community.” Science of the Total Environment 728: 138764 DOI: 10.1016/j.scitotenv.2020.138764.

Ahmed, W., Bertsch, P. M., Bivins, A., Bibby, K., Farkas, K., Gathercole, A., Haramoto, E., Gyawali, P., Korajkic, A., McMinn, B. R., Mueller, J. F., Simpson, S. L., Smith, W. J. M., Symonds, E. M., Thomas, K. V., Verhagen, R. and Kitajima, M. (2020). “Comparison of virus concentration methods for the RT-qPCR-based recovery of murine hepatitis virus, a surrogate for SARS-CoV-2 from untreated wastewater.” Science of the Total Environment 739: 139960 DOI: 10.1016/j.scitotenv.2020.139960.

Ai, Y., Davis, A., Jones, D., Lemeshow, S., Tu, H., He, F., Ru, P., Pan, X., Bohrerova, Z. and Lee, J. (2021). “Wastewater SARS-CoV-2 monitoring as a community-level COVID-19 trend tracker and variants in Ohio, United States.” Science of the Total Environment 801: 149757 DOI: 10.1016/j.scitotenv.2021.149757.

Ash, K., Alamilla, I., Li, Y., Joyner, D., Williams, D., McKay, P., Green, B., Iler, C., DeBlander, S. and Kara-Murdoch, F. (2021). “Coding-Complete Genome Sequence of a SARS-CoV-2 Variant Obtained from Raw Sewage at the University of Tennessee—Knoxville Campus.” Microbiology Resource Announcements 10(47): e01049–01021 DOI: https://doi-org.utk.idm.oclc.org/10.1128/MRA.01049-21.

Ash, K., Li, Y., Alamilla, I., Joyner, D., Williams, D. E., McKay, P., Green, B., Iler, C., DeBlander, S. and North, C. (2023). “SARS-CoV-2 raw wastewater surveillance from student residences on an urban university campus.” Frontiers in Microbiology 14 DOI: 10.3389/fmicb.2023.1101205.

Benefield, A. E., Skrip, L. A., Clement, A., Althouse, R. A., Chang, S. and Althouse, B. M. (2020). “SARS-CoV-2 viral load peaks prior to symptom onset: a systematic review and individual-pooled analysis of coronavirus viral load from 66 studies.” medRxiv: 2020.2009. 2028.20202028 DOI: 10.1101/2020.09.28.20202028.

Bitter, L. C., Kibbee, R., Jiménez, G. C. and Örmeci, B. (2022). “Wastewater Surveillance of SARS-CoV-2 at a Canadian University Campus and the Impact of Wastewater Characteristics on Viral RNA Detection.” Acs Es&T Water 2(11): 2034–2046 DOI: https://10.1021/acsestwater.2c00060.

Bustin, S. A., Benes, V., Garson, J. A., Hellemans, J., Huggett, J., Kubista, M., Mueller, R., Nolan, T., Pfaffl, M. W., Shipley, G. L., Vandesompele, J. and Wittwer, C. T. (2009). “The MIQE Guidelines: Minimum Information for Publication of Quantitative Real-Time PCR Experiments.” Clinical Chemistry 55(4): 611–622 DOI: https://doi-org.utk.idm.oclc.org/10.1373/clinchem.2008.112797.

Collaborative, C.-W. (2023, Auguest 31, 2023). “COVID-19WBECollaborative.“. www.COVID19wbec.org.

D’Aoust, P. M., Mercier, E., Montpetit, D., Jia, J.-J., Alexandrov, I., Neault, N., Baig, A. T., Mayne, J., Zhang, X., Alain, T., Langlois, M.-A., Servos, M. R., MacKenzie, M., Figeys, D., MacKenzie, A. E., Graber, T. E. and Delatolla, R. (2021). “Quantitative analysis of SARS-CoV-2 RNA from wastewater solids in communities with low COVID-19 incidence and prevalence.” Water Research 188: 116560 DOI: 10.1016/j.watres.2020.116560.

Daou, M., Kannout, H., Khalili, M., Almarei, M., Alhashami, M., Alhalwachi, Z., Alshamsi, F., Tahseen Al Bataineh, M., Azzam Kayasseh, M. and Al Khajeh, A. (2022). “Analysis of SARS-CoV-2 viral loads in stool samples and nasopharyngeal swabs from COVID-19 patients in the United Arab Emirates.” PLOS ONE 17(9): e0274961 DOI: 10.1371/journal.pone.0274961.

Feng, S., Roguet, A., McClary-Gutierrez, J. S., Newton, R. J., Kloczko, N., Meiman, J. G. and McLellan, S. L. (2021). “Evaluation of Sampling, Analysis, and Normalization Methods for SARS-CoV-2 Concentrations in Wastewater to Assess COVID-19 Burdens in Wisconsin Communities.” Acs Es&T Water 1(8): 1955–1965 DOI: https://10.1021/acsestwater.1c00160.

Gerrity, D., Papp, K., Stoker, M., Sims, A. and Frehner, W. (2021). “Early-pandemic wastewater surveillance of SARS-CoV-2 in Southern Nevada: Methodology, occurrence, and incidence/prevalence considerations.” Water research X 10: 100086 DOI: 10.1016/j.wroa.2020.100086.

Greenwald, H. D., Kennedy, L. C., Hinkle, A., Whitney, O. N., Fan, V. B., Crits-Christoph, A., Harris-Lovett, S., Flamholz, A. I., Al-Shayeb, B., Liao, L. D., Beyers, M., Brown, D., Chakrabarti, A. R., Dow, J., Frost, D., Koekemoer, M., Lynch, C., Sarkar, P., White, E., Kantor, R. and Nelson, K. L. (2021). “Tools for interpretation of wastewater SARS-CoV-2 temporal and spatial trends demonstrated with data collected in the San Francisco Bay Area.” Water research X 12: 100111 DOI: 10.1016/j.wroa.2021.100111.

Hamouda, M., Mustafa, F., Maraqa, M., Rizvi, T. and Aly Hassan, A. (2021). “Wastewater surveillance for SARS-CoV-2: Lessons learnt from recent studies to define future applications.” Science of the Total Environment 759: 143493 DOI: 10.1016/j.scitotenv.2020.143493.

Jafferali, M. H., Khatami, K., Atasoy, M., Birgersson, M., Williams, C. and Cetecioglu, Z. (2021). “Benchmarking virus concentration methods for quantification of SARS-CoV-2 in raw wastewater.” Science of the Total Environment 755: 142939 DOI: 10.1016/j.scitotenv.2020.142939.

Karthikeyan, S., Ronquillo, N., Belda-Ferre, P., Alvarado, D., Javidi, T., Longhurst, C. A. and Knight, R. (2021). “High-throughput wastewater SARS-CoV-2 detection enables forecasting of community infection dynamics in San Diego County.” Msystems 6(2): 10.1128/msystems.00045-00021 DOI: https://doi-org.utk.idm.oclc.org/10.1128/msystems.00045-21.

Kitajima, M., Sassi, H. P. and Torrey, J. R. (2018). “Pepper mild mottle virus as a water quality indicator.” NPJ Clean Water 1(1): 19 DOI: https://10.1038/s41545-018-0019-5.

Li, X., Zhang, S., Shi, J., Luby, S. P. and Jiang, G. (2021). “Uncertainties in estimating SARS-CoV-2 prevalence by wastewater-based epidemiology.” Chemical Engineering Journal 415: 129039 DOI: 10.1016/j.cej.2021.129039.

Li, Y., Ash, K. T., Joyner, D. C., Williams, D. E., Alamilla, I., McKay, P., Iler, C., Green, B., Kara-Murdoch, F. and Swift, C. M. (2023). “Decay of enveloped SARS-CoV-2 and non-enveloped PMMoV RNA in raw sewage from university dormitories.” Frontiers in Microbiology 14: 1144026 DOI: 10.3389/fmicb.2023.1144026.

Li, Y., Ash, K. T., Joyner, D. C., Williams, D. E., Alamilla, I., McKay, P. J., Iler, C. and Hazen, T. C. (2023). “Evaluating various composite sampling modes for detecting pathogenic SARS-CoV-2 virus in raw sewage.” Frontiers in Microbiology 14 DOI: 10.3389/fmicb.2023.1305967.

Maal-Bared, R., Qiu, Y., Li, Q., Gao, T., Hrudey, S. E., Bhavanam, S., Ruecker, N. J., Ellehoj, E., Lee, B. E. and Pang, X. (2023). “Does normalization of SARS-CoV-2 concentrations by Pepper Mild Mottle Virus improve correlations and lead time between wastewater surveillance and clinical data in Alberta (Canada): comparing twelve SARS-CoV-2 normalization approaches.” Science of the Total Environment 856: 158964 DOI: 10.1016/j.scitotenv.2022.158964.

Medema, G., Heijnen, L., Elsinga, G., Italiaander, R. and Brouwer, A. (2020). “Presence of SARS-Coronavirus-2 RNA in Sewage and Correlation with Reported COVID-19 Prevalence in the Early Stage of the Epidemic in The Netherlands.” Environmental Science & Technology Letters 7(7): 511–516 DOI: https://10.1021/acs.estlett.0c00357.

Nagarkar, M., Keely, S. P., Jahne, M., Wheaton, E., Hart, C., Smith, B., Garland, J., Varughese, E. A., Braam, A., Wiechman, B., Morris, B. and Brinkman, N. E. (2022). “SARS-CoV-2 monitoring at three sewersheds of different scales and complexity demonstrates distinctive relationships between wastewater measurements and COVID-19 case data.” Science of the Total Environment 816: 151534 DOI: 10.1016/j.scitotenv.2021.151534.

Nemudryi, A., Nemudraia, A., Wiegand, T., Surya, K., Buyukyoruk, M., Cicha, C., Vanderwood, K. K., Wilkinson, R. and Wiedenheft, B. (2020). “Temporal Detection and Phylogenetic Assessment of SARS-CoV-2 in Municipal Wastewater.” Cell Reports Medicine 1(6): 100098 DOI: 10.1016/j.xcrm.2020.100098.

Randazzo, W., Truchado, P., Cuevas-Ferrando, E., Simón, P., Allende, A. and Sánchez, G. (2020). “SARS-CoV-2 RNA in wastewater anticipated COVID-19 occurrence in a low prevalence area.” Water Research 181: 115942 DOI: 10.1016/j.watres.2020.115942.

Sapula, S. A., Whittall, J. J., Pandopulos, A. J., Gerber, C. and Venter, H. (2021). “An optimized and robust PEG precipitation method for detection of SARS-CoV-2 in wastewater.” Science of the Total Environment 785: 147270 DOI: 10.1016/j.scitotenv.2021.147270.

Schmitz, B. W., Innes, G. K., Prasek, S. M., Betancourt, W. Q., Stark, E. R., Foster, A. R., Abraham, A. G., Gerba, C. P. and Pepper, I. L. (2021). “Enumerating asymptomatic COVID-19 cases and estimating SARS-CoV-2 fecal shedding rates via wastewater-based epidemiology.” Science of the Total Environment 801: 149794 DOI: 10.1016/j.scitotenv.2021.149794.

Scott, L. C., Aubee, A., Babahaji, L., Vigil, K., Tims, S. and Aw, T. G. (2021). “Targeted wastewater surveillance of SARS-CoV-2 on a university campus for COVID-19 outbreak detection and mitigation.” Environmental Research 200: 111374 DOI: 10.1016/j.envres.2021.111374.

von Sperling, M., Verbyla, M. E. and Oliveira, S. M. A. C. (2020). Relationship between monitoring variables. Correlation and regression analysis. Assessment of Treatment Plant Performance and Water Quality Data: A Guide for Students, Researchers and Practitioners, IWA Publishing: 0.

Walsh, K. A., Jordan, K., Clyne, B., Rohde, D., Drummond, L., Byrne, P., Ahern, S., Carty, P. G., O’Brien, K. K. and O’Murchu, E. (2020). “SARS-CoV-2 detection, viral load and infectivity over the course of an infection.” Journal of Infection 81(3): 357–371 DOI: 10.1016/j.jinf.2020.06.067.

Wolfe, M. K., Archana, A., Catoe, D., Coffman, M. M., Dorevich, S., Graham, K. E., Kim, S., Grijalva, L. M., Roldan-Hernandez, L., Silverman, A. I., Sinnott-Armstrong, N., Vugia, D. J., Yu, A. T., Zambrana, W., Wigginton, K. R. and Boehm, A. B. (2021). “Scaling of SARS-CoV-2 RNA in Settled Solids from Multiple Wastewater Treatment Plants to Compare Incidence Rates of Laboratory-Confirmed COVID-19 in Their Sewersheds.” Environmental Science & Technology Letters 8(5): 398–404 DOI: https://10.1021/acs.estlett.1c00184.

Wu, F., Xiao, A., Zhang, J., Moniz, K., Endo, N., Armas, F., Bonneau, R., Brown, M. A., Bushman, M., Chai, P. R., Duvallet, C., Erickson, T. B., Foppe, K., Ghaeli, N., Gu, X., Hanage, W. P., Huang, K. H., Lee, W. L., Matus, M., McElroy, K. A., Nagler, J., Rhode, S. F., Santillana, M., Tucker, J. A., Wuertz, S., Zhao, S., Thompson, J. and Alm, E. J. (2022). “SARS-CoV-2 RNA concentrations in wastewater foreshadow dynamics and clinical presentation of new COVID-19 cases.” Science of the Total Environment 805: 150121 DOI: 10.1016/j.scitotenv.2021.150121.

Wurtzer, S., Marechal, V., Mouchel, J., Maday, Y., Teyssou, R., Richard, E., Almayrac, J. and Moulin, L. (2020). “Evaluation of lockdown impact on SARS-CoV-2 dynamics through viral genome quantification in Paris wastewaters.” DOI: https://doi-org.utk.idm.oclc.org/10.1101/2020.04.12.20062679

Xiao, A., Wu, F., Bushman, M., Zhang, J., Imakaev, M., Chai, P. R., Duvallet, C., Endo, N., Erickson, T. B., Armas, F., Arnold, B., Chen, H., Chandra, F., Ghaeli, N., Gu, X., Hanage, W. P., Lee, W. L., Matus, M., McElroy, K. A., Moniz, K., Rhode, S. F., Thompson, J. and Alm, E. J. (2022). “Metrics to relate COVID-19 wastewater data to clinical testing dynamics.” Water Research 212: 118070 DOI: 10.1016/j.watres.2022.118070.

Zambrana, W., Catoe, D., Coffman, M. M., Kim, S., Anand, A., Solis, D., Sahoo, M. K., Pinsky, B. A., Bhatt, A. S. and Boehm, A. B. (2022). “SARS-CoV-2 RNA and N antigen quantification via wastewater at the campus level, building cluster level, and individual-building level.” Acs Es&T Water 2(11): 2025–2033 DOI: https://doi-org.utk.idm.oclc.org/10.1021/acsestwater.2c00050.

Zhan, Q., Babler, K. M., Sharkey, M. E., Amirali, A., Beaver, C. C., Boone, M. M., Comerford, S., Cooper, D., Cortizas, E. M., Currall, B. B., Foox, J., Grills, G. S., Kobetz, E., Kumar, N., Laine, J., Lamar, W. E., Mantero, A. M. A., Mason, C. E., Reding, B. D., Robertson, M., Roca, M. A., Ryon, K., Schürer, S. C., Shukla, B. S., Solle, N. S., Stevenson, M., Tallon Jr, J. J., Thomas, C., Thomas, T., Vidović, D., Williams, S. L., Yin, X. and Solo-Gabriele, H. M. (2022). “Relationships between SARS-CoV-2 in Wastewater and COVID-19 Clinical Cases and Hospitalizations, with and without Normalization against Indicators of Human Waste.” Acs Es&T Water 2(11): 1992–2003 DOI: https://10.1021/acsestwater.2c00045.

Zheng, S., Fan, J., Yu, F., Feng, B., Lou, B., Zou, Q., Xie, G., Lin, S., Wang, R. and Yang, X. (2020). “Viral load dynamics and disease severity in patients infected with SARS-CoV-2 in Zhejiang province, China, January-March 2020: retrospective cohort study.” bmj 369 DOI: https://doi-org.utk.idm.oclc.org/10.1136/bmj.m1443

